# Antibody and T cell responses to Sinopharm/BBIBP-CorV in naïve and previously infected individuals in Sri Lanka

**DOI:** 10.1101/2021.07.15.21260621

**Authors:** Chandima Jeewandara, Inoka Sepali Aberathna, Pradeep Darshana Pushpakumara, Achala Kamaladasa, Dinuka Guruge, Deshni Jayathilaka, Banuri Gunasekara, Shyrar Tanussiya, Heshan Kuruppu, Thushali Ranasinghe, Shashika Dayarathne, Osanda Dissanayake, Nayanathara Gamalath, Dinithi Ekanayake, MPDJ Jayamali, Ayesha Wijesinghe, Madushika Dissanayake, Deshan Madusanka, Tibutius Thanesh Jayadas, Anushika Mudunkotuwa, Gayasha Somathilake, Michel Harvie, Thashmi Nimasha, Saubhagya Danasekara, Ruwan Wijayamuni, Lisa Schimanski, Tiong K. Tan, Tao Dong, Alain Townsend, Graham S. Ogg, Gathsaurie Neelika Malavige

## Abstract

**Background:** As there are limited data of the immunogenicity of the Sinopharm/BBIBP-CorV in different populations, antibody responses against different SARS-CoV-2 variants of concern and T cell responses, we investigated the immunogenicity of the vaccine, in individuals in Sri Lanka.

**Methods:** SARS-CoV-2-specific antibodies were measured in 282 individuals who were seronegative at baseline, and ACE2 receptor blocking antibodies, antibodies to the receptor binding domain (RBD) of the wild type (WT), B.1.1.7, B.1.351 and B.1.617.2, ex vivo and cultured IFNγ ELISpot assays, intracellular cytokine secretion assays and B cell ELISpot assays were carried out in a sub cohort of the vaccinees at 4 weeks and at 6 weeks (2 weeks after the second dose).

**Results:** 95% of the vaccinees seroconverted, although the seroconversion rates were significantly lower (p<0.001) in individuals >60 years (93.3%) compared to those who were 20 to 39 years (98.9%). 81.25% had ACE2 receptor blocking antibodies at 6 weeks, and there was no difference in these antibody titres in vaccine sera compared to convalescent sera (p=0.44). Vaccinees had significantly less (p<0.0001) antibodies to the RBD of WT and B.1.1.7, although there was no difference in antibodies to the RBD of B.1.351 and B.1.617.2 compared to convalescent sera. 27.7% of 46.4% of vaccinees had ex vivo IFNγ and cultured ELISpot responses respectively, and IFNγ and CD107a responses were detected by flow cytometry.

**Conclusions:** Sinopharm/BBIBP-CorV appeared to induce high seroconversion rates and induce a similar level of antibody responses against ACE2 receptor, B.1.617.2 and B.1.351 as seen following natural infection.

## Introduction

Although the COVID-19 pandemic has caused unprecedented mortality and morbidity in almost all countries in the world, within a period of one year, several vaccines for COVID-19 have been developed. The mRNA vaccine Pfizer BioNTech (BNT162b2) was the first to be approved by the WHO in December 2020, followed by many other vaccines such as AZD1222 (Astrazeneca), Jansen Ad26, COV2.S, Moderna mRNA1273, Sinopharm/BB1BP-CorV and Sinovac^1^. During this short time, many of these vaccines have been extensively studied, including the persistence of neutralization antibody responses, antibody responses to SARS-CoV-2 variants of concern (VOCs), memory T cell and B cell responses, memory T cell and B cell repertoire^2-4^. However, there are limited data regarding the immune responses of the Sinopharm/BBIBP-CorV in real-world situations, memory B cell and T cell responses and responses to SARS-CoV-2 VOCs.

Sinopharm/BBIBP-CorV is an inactivated vero-cell derived vaccine, which demonstrated high levels of neutralizing antibodies in animal models and prevented infection in animal challenge models^5^. The vaccine was also shown to be well tolerated and 79% to 96% individuals were shown to seroconvert at 14 days after the second dose, while all individuals (100%) were shown to seroconvert by 28 days. All individuals in the vaccine arm were reported to have neutralizing antibodies by 42 days, following the second dose of the vaccine^6^. The phase 3 demonstrated an efficacy of 78.1% against symptomatic illness^7^, and it was given emergency user license by the WHO on 7^th^ May 2021. Sinopharm/BBIBP-CorV is the predominant vaccine used in many Asian and Middle East countries and was recently included in the Global Alliance for Vaccines and Immunizations (GAVI), to be distributed under the COVAX program^8^.

Although safety, immunogenicity and efficacy data for Sinopharm/BBIBP-CorV are available, there are limited data on immunogenicity in different populations, and there are limited data regarding T cell responses to this vaccine in humans or responses to the emerging VOCs such as B.1.617.2 (“delta variant”). The related delta variants have been shown to result in immune evasion and escape neutralization by vaccine induced neutralization antibodies and by convalescent sera, although to a lesser extent than beta (B.1.351)^9^. However, although VOCs have been shown to escape immunity induced by antibodies, they were less likely to evade T cell immunity^10^. Given that the delta is the dominant variant in many countries and that virus specific T cells may play an additional role in protection against severe COVID-19^11^, it would be important to investigate immunogenicity of this vaccine against VOCs and also to assess virus-specific T cell responses.

Sinopharm/BBIBP-CorV is the vaccine most widely used in Sri Lanka and as there are no data regarding its immunogenicity in real world situations and to VOCs, we investigated antibody responses to the SARS-CoV-2 including to the VOCs, along with T cell responses and their functionality, including memory B cell responses in a large cohort of Sri Lankan individuals.

## Methods

### Study participants

323 individuals above the age of 21 years, who were vaccinated in Colombo, Sri Lanka, were included in the study following informed written consent. A baseline blood sample was obtained to determine previous SARS-CoV-2 infection, at 4 weeks when the second dose has been administered and again 2 weeks from obtaining the second dose of the vaccine (6 weeks from first dose). Presence of comorbid illnesses such as diabetes, hypertension and chronic kidney disease was recorded. Thirty-six individuals with varying severity of past COVID-19 were also recruited six weeks from the onset of illness (supplementary methods), to compare the antibody responses for ACE2 receptor blocking assays and the antibody responses to the VOCs.

Ethics approval was obtained from the Ethics Review Committee of the University of Sri Jayewardenepura.

### Detection of SARS-CoV-2 specific antibodies

Seroconversion rates to the BBIBP-CorV vaccine were determined by using the Wantai SARS-CoV-2 Ab ELISA (Beijing Wantai Biological Pharmacy Enterprise, China), which detects IgM, IgG and IgA antibodies to the receptor binding domain (RBD) of the SARS-CoV-2. A cut-off value for each ELISA was calculated according to manufacturer’s instructions. Based on the cut off value, the antibody index (used as an indirect indicator of the antibody titre) was calculated by dividing the absorbance of each sample by the cutoff value, according to the manufacturer’s instructions.

### Surrogate neutralizing antibody test (sVNT) to detect ACE2 receptor blocking antibodies

Due to the non-availability of biosafety level 3 facilities to carry out live neutralization assays a surrogate virus neutralization test (sVNT)^12^, which measures the percentage of inhibition of binding of the RBD of the S protein to recombinant ACE2^12^ (Genscript Biotech, USA) was carried out according the manufacturer’s instructions as previously described by us^13^. Inhibition percentage ≥ 25% in a sample was considered as positive for NAbs.

### Haemagglutination test (HAT) to detect antibodies to the receptor binding domain (RBD)

The HAT was carried out as previously described using the B.1.1.7 (N501Y), B.1.351 (N501Y, E484K, K417N) and B.1.617.2 (L452R, T478K) versions of the IH4-RBD reagents^14^, which included the relevant amino acid changes introduced by site directed mutagenesis^15^. The assays were carried out and interpreted as previously described ^16^. Details of the methods are available in the supplementary methods.

### Ex vivo and cultured IFNγ ELISpot assays and intracellular cytokine staining and B cell ELISpots

Ex vivo IFNγ ELISpot assays were carried out using freshly isolated peripheral blood mononuclear cells (PBMC) obtained from 66 individuals. Details of the ex vivo and cultured ELISpot assays is available in supplementary methods. For ex vivo and cultured IFNγ ELISpot assays, responses to two pools of overlapping peptides named S1 (peptide 1 to 130) and S2 (peptide 131 to 253) covering the whole spike protein (253 overlapping peptides) were assessed.

The expression of CD107a and IFN-g were determined on both CD4 and CD8 T cells by flowcytometry in freshly isolated PBMC as described previously^17^. Cells were acquired on a BD FACSAria III Cell Sorter using DIVA v8 software (BD Biosciences, USA). The frequency of SARS-CoV-2 S1, S2 and N recombinant protein specific memory B cells were assessed using B cell ELISpot assays. All experiments were carried out in duplicate and anti-human IgG monoclonal capture antibodies, was used as a positive control, and media alone as a negative control. Details of the methods used is available in supplementary methods.

### Statistical analysis

The percentage calculations for the analysis were conducted in Microsoft Excel and the 95% confidence intervals for each category were calculated using the R software (version 4.0.3) and R-studio (version 1.4.1106). Pearson Chi Square Association tests were performed at a confidence level of 95% using the R software in order to identify the statistically significant associations of the age categories and the sex of the respondents in the study with the 4 weeks and 6 weeks post-vaccine antibody results. The differences in antibodies at baseline uninfected and infected individuals were assessed by the Wilcoxon matched pairs signed ranked test. All tests were two sided. The differences in the antibody titres between different age groups was determined by the Kruskal-Wallis test.

## Results

Of the 323 individuals, 41 individuals were found to have SARS-CoV-2 antibodies at the time of enrollment to the study (at the time of receiving the first dose). Of the 282 individuals who were seronegative, 111 (39.4%) were females. The distribution of individuals, seropositivity rates at 4 weeks and 6 weeks and the median antibody titers and interquartile ranges, are shown in Table 1. The overall seroconversion rates were 95.0% at 6 weeks. The seroconversion rates were highest in the 20 to 39 age group (98.88%) at 6 weeks (2 weeks after obtaining the 2^nd^ dose) and were significantly different in the three age groups (Pearson Chi-Square = 842.983, p<0.001). The seroconversion rates were also higher in males (96.5%, 95% CI 93.73%, 99.25%) compared to females (92.8%, 95% CI87.98%, 97.60%), which was significantly different (Pearson Chi-Square = 836.750, p <0.001). 48 (17.0%) individuals had comorbidities (diabetes, hypertension of chronic kidney disease). There was no difference in seroconversion rates in those with comorbidities (91.7%, 95 CI, 83.8% to 99.5%) compared to those who did not have comorbidities (95.7%, 95% CI 93.1% to 98.3%).

**Table 1:** Seroconversion rates and SARS-CoV-2 specific total antibody levels (indicated by antibody index) in individuals who received the Sinopharm/BBIBP-CorV vaccine at 4 weeks and 6 weeks.

| Age group | Seropositivity at 4 weeks<br>N (%), 95% confidence interval | Antibody index<br>(antibody titre) at 4 weeks<br>Median, IQR | Seropositivity at 6 weeks<br>N (%) | Antibody index<br>(antibody titre) at 6 weeks<br>Median, IQR |
| --- | --- | --- | --- | --- |
| 21 to 40<br>N=89 | 54 (60.67%)<br>(50.53%, 70.82%) | 1.79 (0.58 to 4.48) | 88 (98.88%)<br>(96.69%, 100%) | 13.22 (12.47 to 13.72) |
| 41 to 60<br>N=163 | 82 (50.31%)<br>(42.63%, 57.98%) | 1.0 (0.13 to 3.85) | 152 (93.25%)<br>(89.40%, 97.10%) | 12.6 (8.09 to 13.52) |
| >60<br>N=30 | 19 (63.33%)<br>(46.09%, 80.58%) | 1.75 (0.55 to 4.77) | 28 (93.33%)<br>(84.41%, 100%) | 12.42 (5.41 to 13.52) |

### ACE2 receptor binding antibodies in baseline naïve and previously infected individuals

ACE2 receptor blocking antibodies were measured in vaccinees who were baseline non-infected (n=64) and baseline infected (n=38) and in naturally infected individuals (n=36). A percentage of inhibition of 25% was considered as a positive response as previously described^13^. At 4 weeks, 24/64 (37.5%) of uninfected individuals had a positive response, while 52/64 (81.25%) had a response at 6 weeks (Figure 1B). 32/36 (88.9%) of naturally infected individuals had a positive response. The antibody levels significantly increased from baseline (median=5.2, IQR 0.3 to 8.5 % of inhibition) to 4 weeks (median=20.96, IQR=14.9 to 34.4, p<0.0001 % of inhibition) and from 4 weeks to 6 weeks (median= 78.7, IQR 44.1 to 91.2, p<0.0001), in non-infected individuals. The median ACE2 receptor blocking antibodies of those who were naturally infected was 66.7 % (IQR 44.2 to 84.9 % of inhibition). There was no significant difference in antibody levels in those who were naturally infected compared to these vaccinees at the end of 6 weeks (p=0.15). There was no correlation with age and the ACE2 receptor blocking antibodies at 4 weeks (Spearman’s r=0.04, p=0.69) or at 6 weeks (Spearman’s r=-0.21, p=0.10) and there was no difference in the antibody titres of the three different age groups at 4 weeks or at 6 weeks (supplementary figure 1).

**Figure 1:**
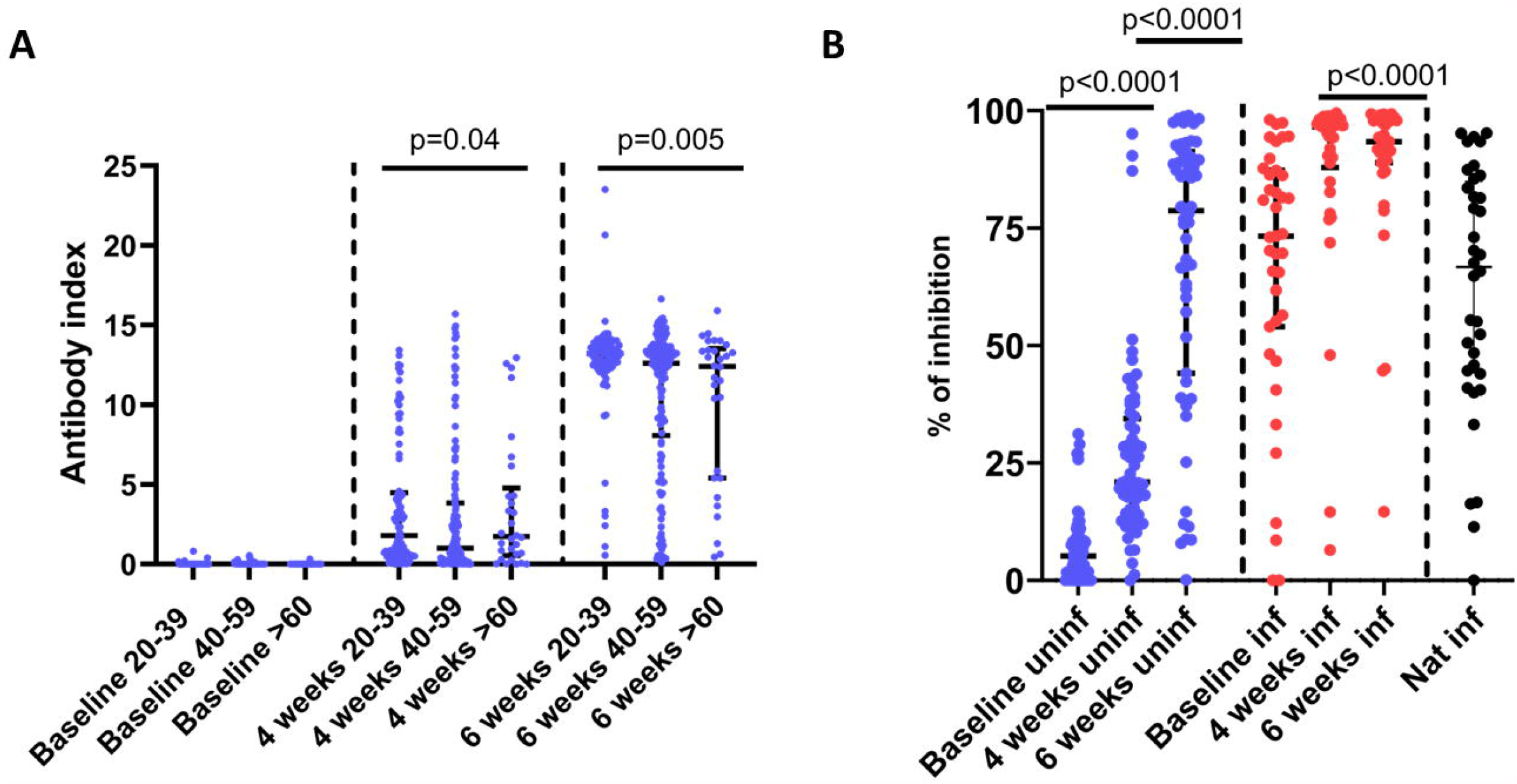
SARS-CoV-2 specific antibody responses (total antibodies and ACE2 receptor blocking antibodies in those who received the Sinopharm.BBIBP-CorV vaccine. SARS-CoV-2 specific total antibodies were measured by ELISA in 20- to 39-year-old individuals (n=89), 40- to 59-year-olds (n=163) and >60 years olds (n=30) at 4 weeks following the first dose and again at 6 weeks (2 weeks after the second dose) (A). ACE2 receptor blocking antibodies were measured by the surrogate neutralizing antibody assay in a sub cohort of these individuals (n=64) at 4 weeks and at 6 weeks, in a cohort of individuals (n=38), who were found to be seropositive for SARS-CoV-2 at the time of recruitment and in a cohort of naturally infected individuals 6 weeks following onset of illness (n=36) (B). The differences in the total antibody titres between different age groups was determined by the Kruskal-Wallis test. The differences in ACE2 receptor blocking antibodies at baseline uninfected and infected individuals were assessed by the Wilcoxon matched-pairs signed ranked test. All tests were two sided. The error bars indicate the median and the interquartile ranges.

Of those who were previously infected (seropositive at baseline), 4/38 (10.5%) did not have ACE2 blocking antibodies. By 6 weeks, except for one individual, all others had detectable antibodies (Figure 1B). Although the antibody levels significantly increased from baseline to 4 weeks (p<0.0001), there was no significant increase from 4 to 6 weeks (p=0.44).

### HAT assay to determine antibodies to the RBD of the wild type and SARS-CoV-2 variants

Antibodies to the RBD were measured by HAT to the wild type (WT), B.1.1.7, B.1.351 and B.1.617.2 at 4 weeks (n=62) and at 6 weeks (n=58) in individuals who were seronegative for SARS-CoV-2 at time of recruitment. The same cohort of individuals were used in for measuring ACE2 receptor blocking antibodies, HAT, T cell and B cell responses. There was no difference between the HAT titres to the WT strain and B.1.1.7 (p=0.1) at 4 weeks, whereas antibody levels were significantly lower to B.1.617.2 (p=0.04) and B.1.351 (p<0.0001) (Figure 2A). At 6 weeks (2 weeks after the second dose), the antibody responses were significantly lower (p<0.0001) for B.1.1.7, B.1.351 and B.617.2 compared to responses to the WT. A HAT titre of 1:20 was considered as a positive result to the RBD by the HAT assay, as previously described^18^. At 4 weeks, 20/62 (32.2%) had a positive response to the WT, 22/62 (35.4%) to B.1.1.7, 3/62 (4.8%) to B.1.351 and 19/62 (30.6%) to B.1.617.2. At 6 weeks (two weeks after the second dose), 51/58 (87.9%) had a positive response to the WT, 50/58 (86.2%) to B.1.1.7, 16/58 (27.6%) for B.1.351 and 50/58 (86.2%) to B.1.617.2. At 6 weeks, there was a 1.3-fold reduction in the geometric means of the antibody titres to B.1.1.7 (mean 132.4, SD±254.8) compared to the WT ((mean 169.3, SD±258.4) 10.01-fold reduction to B.1.351 (mean 16.9, SD±48.5) and a 1.38-fold reduction to B.617.2 (mean 122.1, SD±218.1). B.1.617.2 had a 1.1-fold reduction in antibodies to RBD, compared to B.1.1.7. There were no significant differences in the HAT titres to the WT, or variants between the different age groups at 4 weeks since receiving the first dose and at 6 weeks (supplementary figure 2).

**Figure 2:**
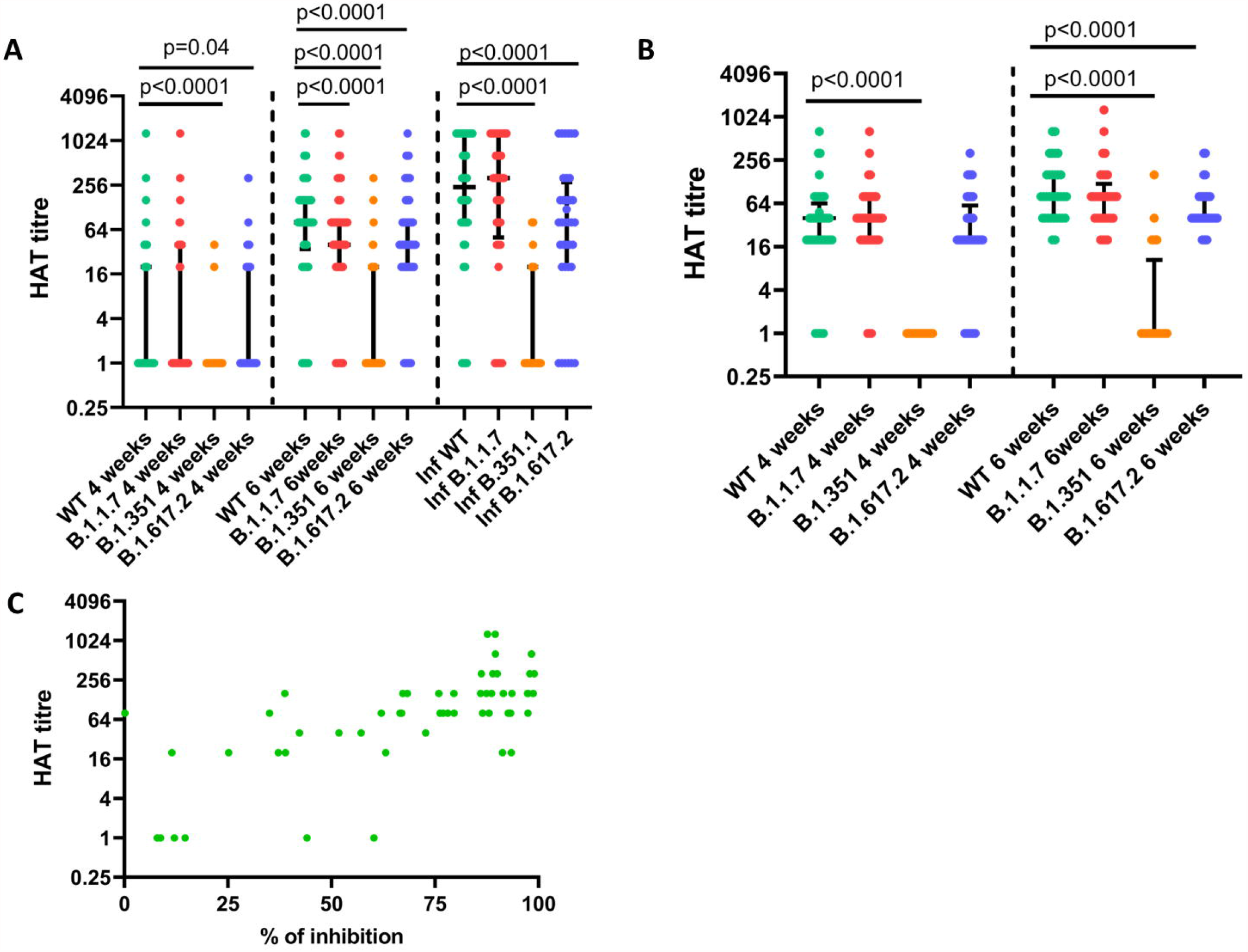
Antibodies to the RBD of SARS-CoV-2 Wuhan (WT) virus and the variants of concern by the haemagglutination test (HAT). Antibodies to the RBD were measured by HAT in previously uninfected individuals at 4 weeks (n=62) and at 6 weeks (n=58), and in individuals who were naturally infected (n=36) for the WT, B.1.1.7, B.351.1 and B.1.617.2 (A). Antibodies to the RBD were also measured by HAT in previously infected individuals (n=33), at 4 weeks and 6 weeks to WT, B.1.1.7, B.1.351 and B.1.617.2 (B). The ACE2 receptor blocking antibody titres, were correlated with the HAT titre to the WT (Spearman’s r=0.67, p<0.0001).The differences in HAT titres to WT and variants in baseline uninfected and infected individuals were assessed by the Wilcoxon matched pairs signed ranked test. All tests were two sided. The error bars indicate the median and the interquartile ranges.

In order to compare the responses to the RBD of vaccinees with responses of convalescent sera, we analyzed the antibodies to RBD in 36 individuals who had varying severity of natural infection. Their onset of illness was 6 weeks before recruitment to the study (same time point as the second bleed of the vaccinees). We found that those with natural infection had significantly less (p<0.0001) HAT titres to B.1.351 and B.1.617.2 than the WT but not B.1.1.7 (Figure 2A). Those who were naturally infected had significantly higher HAT titres to the WT (p=0.005) and B.1.1.7 (p=0.0002), than those who received the vaccine at 6 months. However, there was no significant difference for the HAT titres between those who were naturally infected, compared to the vaccinees for B.1.351 (p=0.89) and B.1.617.2 (p=0.13). Those who were naturally infected had 3.1-fold higher geometric means of antibodies to RBD (HAT titres) to the WT, 3.9-fold higher titres to B.1.1.7, no difference to B.1.351 and 2.3-fold higher titres to B.1.617.2.

Antibody responses were also assessed in baseline infected vaccinees at 4 weeks (n=41) and at 6 weeks (n=33, two weeks after the second dose). There was no significant difference in HAT titres to B.1.1.7 (p=0.30), and B.1.617.2 (p=0.44), compared to the WT, but the levels were significantly lower to B.1.351 (p<0.0001) at 4 weeks (Figure 2B). At 6 weeks, again there was no difference between the titres of WT and B.1.1.7 (p=0.15) but was significantly lower for B.1.351 (p<0.0001) and B.1.617.2 (p<0.0001). The HAT titres significantly increased to WT (p=0.0004), B.1.1.7 (p=0.02), B.1.351 (p=0.008) and B.1.617.2 (p=0.003), following the second dose of the vaccine. After 1 dose, at 4 weeks, 38/41 (92.7%) had a positive response to WT, 38/41 (92.7%) to B.1.1.7, 0/41 (0%) to B.1.351 and 33/41 (80.5%) to B.1.617.2. By 6 weeks, all 33 individuals had a positive response to the WT, B.1.1.7 and B.1.617.2, but only 8/33 (24.2%) had a positive response to B.1.351 (Figure 2B).

The ACE2 receptor blocking antibody titre, significantly correlated with the HAT titre to the WT (Spearman’s r=0.67, p<0.0001) (Figure 2C), B.1.1.7 (Spearman’s r=0.67, p<0.0001), B.1.351 (Spearman’s r=0.38, p=0.004) and B.1.617.2 (Spearman’s r=0.76, p<0.0001) (data not shown).

### T cell responses to the Sinopharm/BBIBP-CorV

We investigated ex vivo IFNγ ELISpot responses in 66 individuals at 4 weeks and at 6-week (n=58). As a positive response was defined as mean±2 SD of the background responses, a cut-off, of 160 spot forming units (SFU) was considered as the threshold response. Accordingly at 4 weeks, 5/66 had responses to the S1 pool of peptides, and 5/66 had responses to S2 (Figure 3A). At 6 weeks, 18/66 (27.7%) had a positive response to S1, while 5/58 had responses to S2. The S1 specific responses at 4 weeks (median 20, IQR 0 to 45 SFU/1 million PBMCs) significantly increased (p<0.0001) by 6 weeks (median 95, IQR 35 to 207.5 SFU/1 million PBMCs). However, there was no difference (p=0.09) in responses to S2 at 4 weeks (median 5, IQR 0 to 43.7 SFU/1 million PBMCs) compared to those at 6 weeks (median 55, IQR 15 to 92.5 SFU/1 million PBMCs). There was no correlation of ex vivo IFNγ ELISpot responses to S1 or S2 or the total S, with age, at 4 weeks or at 6 weeks or with ACE2 receptor blocking antibodies or HAT RBD titres.

**Figure 3:**
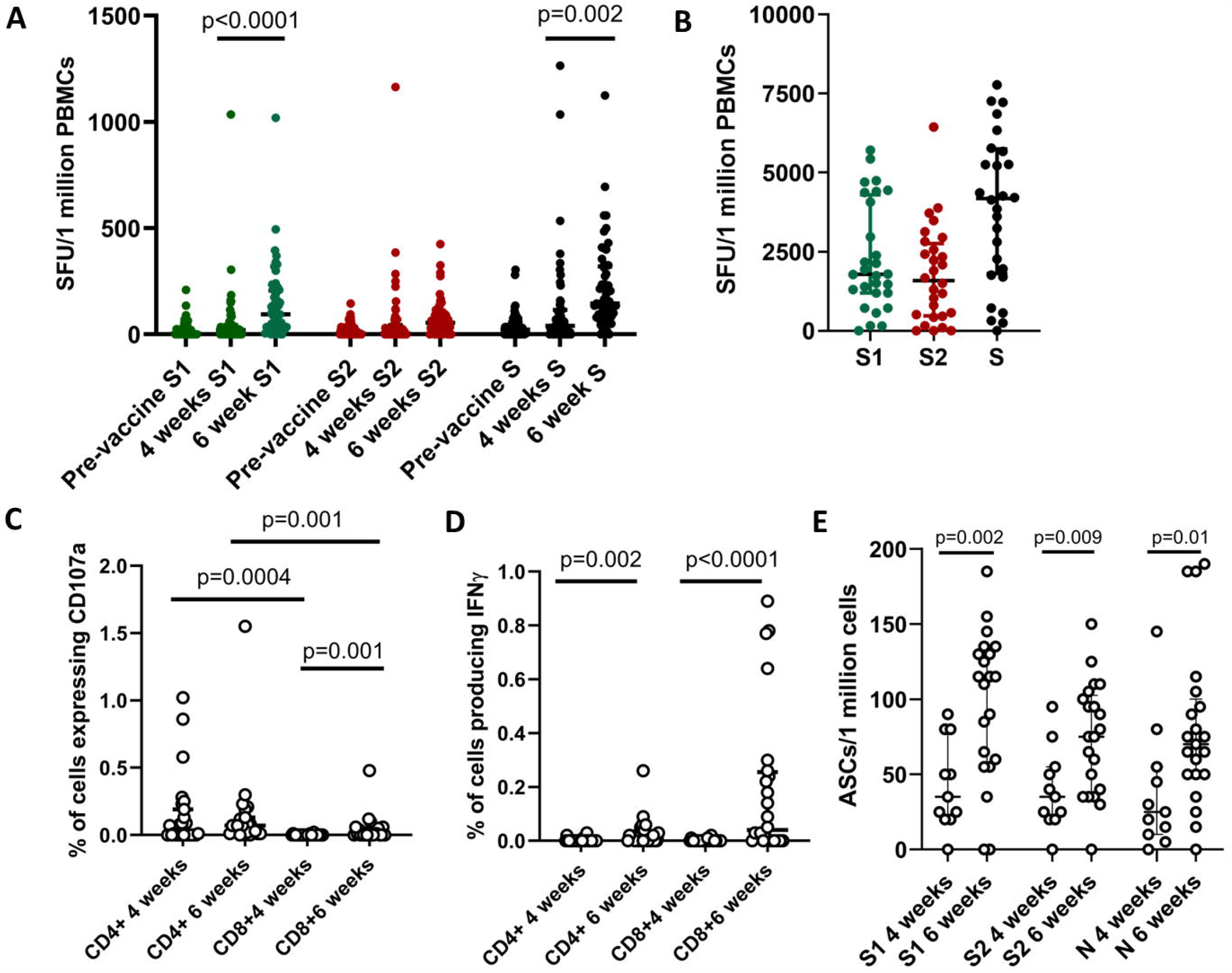
SARS-CoV-2 specific T cell and B cell responses in individuals who received the Sinopharm.BBIBP-CorV vaccine. Ex vivo IFNγ ELISpot assays were carried out at 4 weeks (n=66) and 6 weeks (n=58, two weeks after the second dose) for overlapping peptides of spike protein, which were in two pools S1 and S2 (A). Cultured IFNγ ELISpot assays were carried out at 6 weeks (n=28), using the same pools (B). Intracellular cytokine staining was used to determine CD107a expression (C) and IFNγ production (D) at 4 weeks (n=27) and at 6 weeks (n=23) in CD4+ and CD8+ by flowcytometry. The number of antibody secreting B cells (ASCs) were determined at 4 weeks (n=11) and at 6 weeks (n=21) by B cell ELISpot assays. Wilcoxon matched pairs signed ranked test was used to find out the differences in ex vivo and cultured ELISpot responses for S1 and S2, and differences in CD107a expression and IFNγ production by CD4+ and CD8+ T cells at 4 weeks and 6 weeks. All tests were two sided. The error bars indicate the median and the interquartile ranges.

Cultured IFNγ ELISpot assays were carried out at 6 weeks (2 weeks since obtaining the second dose) in 28 individuals (only 28 were included due to limitation in cell numbers). As a positive response was defined as mean±2 SD of the background responses, a cut off, of 1620 SFU was considered as the response threshold. Accordingly, 13/28 had responses to S1, 11/28 had responses to S2 (Figure 3B). Although responses to S1 pool of peptides was higher than for S2, this was not significant (p=0.09). There was no correlation between ex vivo and cultured IFNγ ELISpot responses at 6 weeks (Spearman’s r=0.15, p=0.44).

Intracellular cytokine staining (ICS) to assess CD107a expression and IFNγ production by CD4+ and CD8+ T cells was carried out at 4 weeks (n=27) and at 6 weeks (n=23) for the overlapping peptides of the spike protein. The CD4+ T cells demonstrated increased expression of CD107a than CD8+ T cells at 4 weeks (p=0.0004) and 6 weeks (p=0.001) (Figure 3C). Although, there was no significant difference in expression of CD107a from 4 weeks to 6 weeks for CD4+ T cells (p=0.58) the expression significantly increased in CD8+ T cells (p=0.002). At 4 weeks since the first dose, there was no IFNγ production detected by CD4+ T cells, and only CD8+ T cells of two individuals produced IFNγ (Figure 3D). IFNγ production by CD4+ T cells (p=0.002) and CD8+ T cells (p<0.0001), significantly increased from 4 weeks to 6 weeks. In contrast to CD107a expression, CD8+ T cells (median 0.04, IQR 0 to 0.25 % of CD8+ T cells) produced significantly more (p=0.02) IFNγ at 6 weeks than CD4+ T cells (median 0.01, IQR 0 to 0.03 % of CD4+ T cells).

### Memory B cell responses to Sinopharm/BB1BP-CorV

Antibody secreting memory B cell responses were assessed by B cell ELISpot assays in 4 weeks (n=11) and at 6 weeks (n=21). A positive response was defined as mean±2 SD of the background responses. Accordingly, a cut-off of 62.9 antibody secreting cells (ASCs)/1 million cells was considered as the positive threshold for S1 protein, 35.7 for S2 and 47.7 for N at 4 weeks and 29.5 for S1, 38.8 for S2 and 35.4 ASCs/1 million cells for N at 6 weeks. At 4 weeks, 3/11 individuals had responses to S1, 5/11 for S2 and 2/11 for N. At 6 weeks, 19/21 had responses to S1, 16/21 for S2 and 17/21 for N protein. The ASC responses significantly increased from 4 weeks to 6 weeks for S1 (p=0.002), S2 (p=0.009) and N (p=0.01) (Figure 3E).

## Discussion

In this study we have investigated the SARS-CoV-2 specific total antibodies, ACE2 receptor blocking antibodies, antibody responses to the RBD of SARS-CoV-2 VOCs, ex vivo and memory T cell responses, T cell functionality and memory B cell responses, in Sri Lankan individuals following the Sinopharm/BBIBP-CorV. As individuals are considered fully vaccinated, two weeks after obtaining the second dose of a COVID-19 vaccines (for those which administer two doses)^19^, we investigated the immune responses at 4 weeks after the first dose and two weeks after obtaining the second dose. We found that two weeks after obtaining the second dose of the vaccine, 95% of individuals seroconverted, although seroconversion rates were significantly lower in those who were >60 years of age (93.3%), compared to those in the 20 to 39 age group (98.9%). The seroconversion rates in this cohort were higher than reported in the phase 1 and 2 trials, probably as our cohort was larger^6^.

Although we could not assess the neutralizing antibodies in our cohort, we used a surrogate neutralizing test, which has shown to correlate well with neutralizing antibodies^12^. We found that by 6 weeks (2 weeks following the second dose), 81.25% of individuals had ACE2 receptor blocking antibodies, which was lower than reported in the phase 1 and II trials, which used live virus assays^6^. However, the ACE2 receptor blocking antibody titres 2 weeks after the second dose of the vaccine, was similar to the levels seen in convalescent sera. Therefore, the vaccine appears to induce a similar level of ACE2 receptor blocking antibodies as following natural infection.

It was shown that both convalescent sera and sera from individuals who received the Sinopharm/BBIBP-CorV vaccine had reduced neutralizing capacity of B.1.617.2 and to a greater degree to B.1.351^20^. The vaccinees had a significant reduction of RBD binding antibodies to the VOCs compared to the WT, and the vaccinees’ HAT titres to the RBD was significantly lower to the WT and the B.1.1.7. However, there was no significant difference in the HAT titres for B.1.617.2 and B.1.351 in the vaccinees when compared to those who were naturally infected, suggesting that the vaccinees had a similar level of protection against infection with delta and beta, as those who were naturally infected. The vaccinees only had a 1.38-fold reduction in the RBD binding antibodies to B.1.617.2 compared to the WT, whereas a 10-fold reduction was seen against B.1.351, suggesting vaccinees are likely to have higher efficacy to B.1.617.2 than for B.1351. Since the reduction in antibodies to RBD was 1.38, compared to the WT virus, the reduction in vaccine efficacy is likely to be less in countries where delta is the predominant variant, which would be important to assess by carefully conducted surveillance using Sinopharm/BBIBP-CorV.

Data regarding T cell and B cell responses to this vaccine are not available. We found that 27.7% of individuals had ex vivo IFNγ ELISpot responses in high frequency to the spike protein overlapping peptides. However, the magnitude of ex vivo IFNγ ELISpot responses and the number of individuals who had responses were less than for a single dose of the AZD1222 vaccine^16^. The AZD1222, has shown to induce high frequency and magnitude of polyfunctional T cell responses even after a single dose of the vaccine^3,21^. We found that Sinopharm/BBIBP-CorV induced potent IFNγ responses as detected by ICS from CD8+ T cells, 2 weeks following the second dose, although degranulation (CD107a) responses predominantly seen from the CD4+ T cell subset. The CD107a expression by CD8+ T cells after the 2^nd^ dose of Sinopharm/BBIBP-CorV appear to be similar to those observed after a single dose of the AZD1222^3^.

In summary, 95% of individuals appear to seroconvert following the Sinopharm/BBIBP-CorV vaccine, and the vaccine appears to induce similar levels of ACE2 receptor blocking antibodies and RBD binding antibodies to B.1.617.2 as seen following natural infection. However, seroconversion rates and immunogenicity appear to be lower in older individuals. The vaccine induced robust T cell responses and memory B cell responses in the vaccinees although the magnitude of responses were less than those observed with some other vaccines.

## Data Availability

All data is available in the manuscript, figures and all supporting information files.

## Acknowledgement

We are grateful to the World Health Organization, UK Medical Research Council and the Foreign and Commonwealth Office for support. T.K.T. is funded by the Townsend-Jeantet Charitable Trust (charity number 1011770) and the EPA Cephalosporin Early Career Researcher Fund. A.T. are funded by the Chinese Academy of Medical Sciences (CAMS) Innovation Fund for Medical Science (CIFMS), China (grant no. 2018-I2M-2-002).

## Competing interests

None of the authors have any conflicts of interest.

## Notes

### Competing Interest Statement

The authors have declared no competing interest.

### Author Declarations

Ethics approval was obtained from the Ethics Review Committee of the University of Sri Jayewardenepura, Sri Lanka.

