## Supplementary figures for "Antibody and T cell responses to Sinopharm/BBIBP-CorV in naïve and previously infected individuals in Sri Lanka"

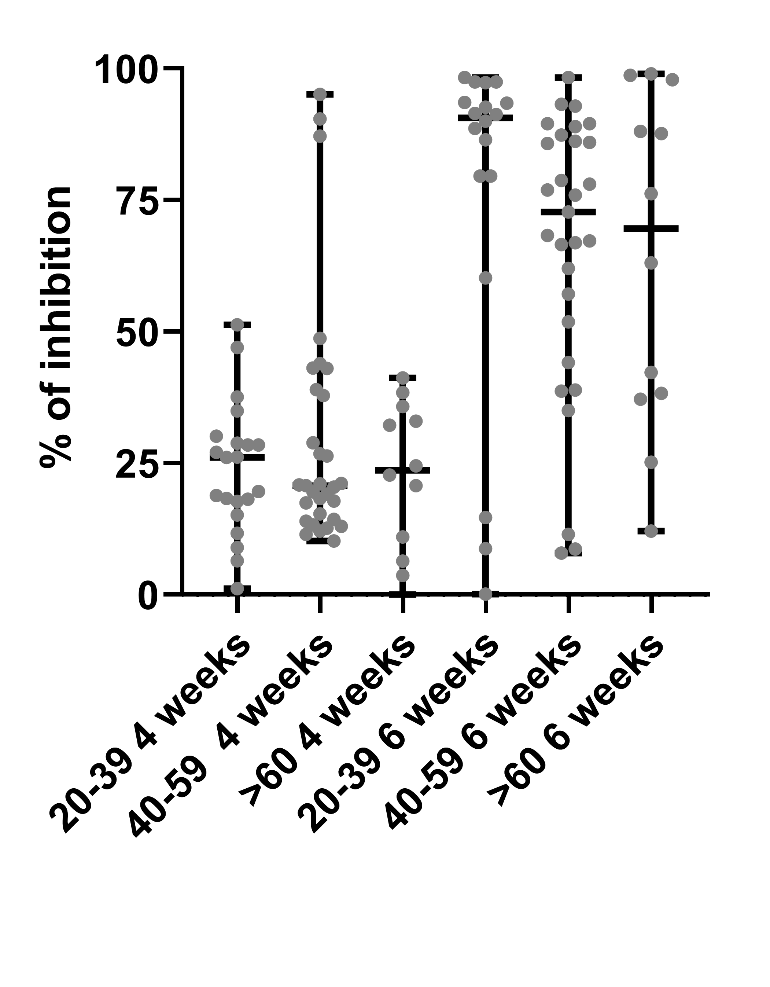

**Supplementary figure 1:** **SARS-CoV-2 CE2 receptor blocking antibodies in those who received the Sinopharm.BBIBP-CorV vaccine.** ACE2 receptor blocking antibodies were measured by the surrogate neutralizing antibody assay in individuals who were 20 to 39 (n=21), 40 to 59 (n=31) and >60 (n=12) at 4 weeks and at 6 weeks. The differences in the total antibody titres between different age groups was determined by the Kruskal-Wallis test. All tests were two sided. The error bars indicate the median and the interquartile ranges.

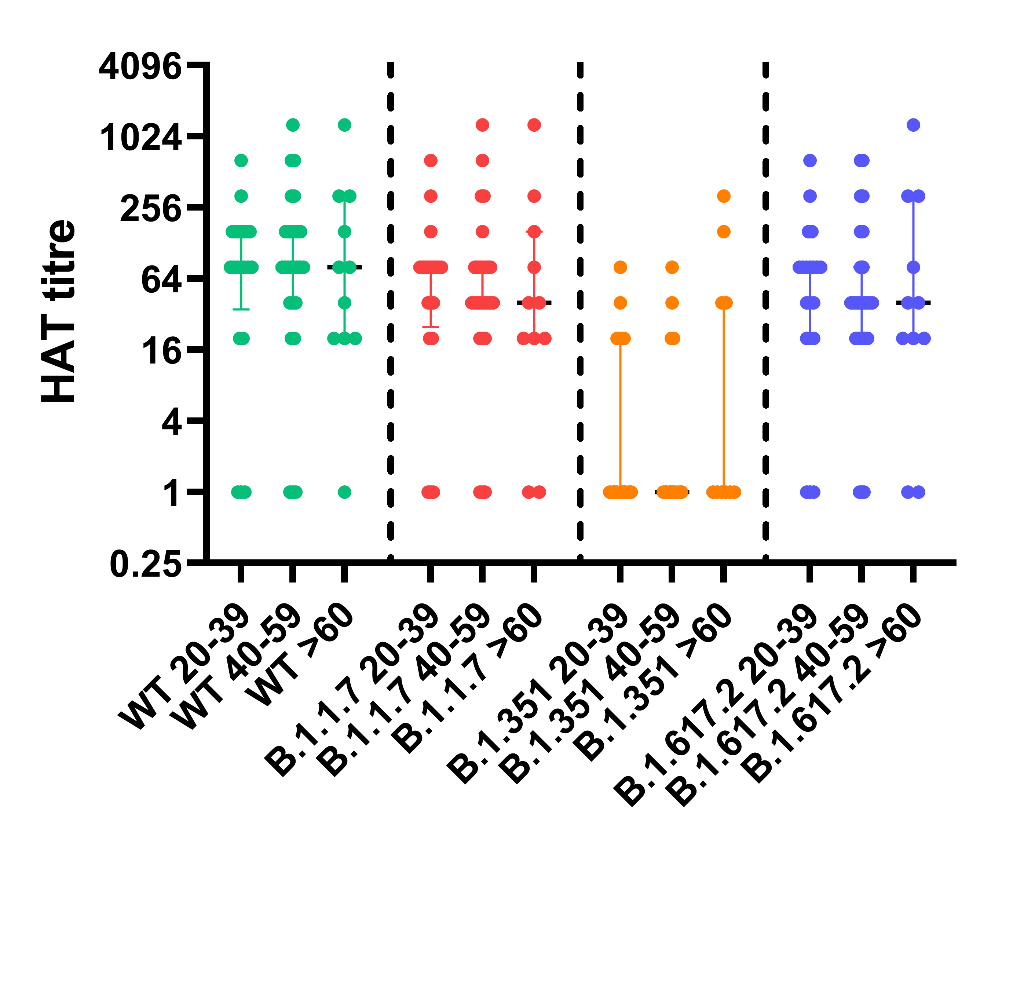

**Supplementary figure 2: Antibodies to the RBD of SARS-CoV-2 Wuhan (WT) virus and the variants of concern by the haemagglutination test (HAT) in individuals of different age groups.** Antibodies to the RBD were measured by HAT for WT, B.1.1.7, B.351.1 and B.1.617.2 in previously uninfected individuals who were 20 to 39 (n=20), 40 to 59 (n=27) and >60 (n=11) at 6 weeks (2 weeks since receiving the second dose of the vaccine). The differences in the HAT titres between different age groups was determined by the Kruskal-Wallis test. All tests were two sided. The error bars indicate the median and the interquartile ranges.
