## Supplementary methods for "Antibody and T cell responses to Sinopharm/BBIBP-CorV in naïve and previously infected individuals in Sri Lanka"

**Patient characteristics**

Twenty patients confirmed with SARS-CoV2 infection based on the positive RT-PCR who were treated at a COVID-19 treatment hospital, were recruited 6 weeks after onset of illness, following informed written consent. Severity of clinical illness and details of laboratory test results were retrieved from the diagnosis card issued by hospitals when the patients were discharged from hospital. Clinical disease severity was classified as mild, moderate and severe according to the WHO guidance on COVID-19 disease severity^1^. Based on the WHO COVID-19 disease classification, 32 patients had mild illness and 4 patients had moderate/severe illness.

**Sample size calculation**

Proportionate stratified sampling was used in order to obtain a sample with a representative mix of respondents from each age category to assess seroconversion rates, as it ensures that each subgroup of a given population is adequately represented within the whole sample population of the research study. An overall minimum sample size of 384 was calculated by the Cochran's Formula with a 5% margin of error, a 95% confidence interval and a 50% response rate as shown below.

Cochran’s sample size calculation formula	n = [z^2^ p (1-p)] / c^2^	where,

n = estimated minimum sample size for an unknown population size

z = standard normal deviation set at 95% confidence level = 1.96

p = percentage picking a choice or response = 0.5

e = margin of error (desired level of precision) = 0.05

n = [1.96^2^ x 0.5 x (1-0.5)] / 0.05^2^ = 384

After estimating the overall sample size, we used convenience sampling to proportionately estimate the required sample sizes from each of the five strata considered for the study. The estimated values for the seroconversion rates are as given below.

The following numbers were resulted after calculation with the proportionately estimated percentages with a sample size of 384.

20-30 years: 7% = 26

30-40 years: 28% = 108

40-50 years: 28% = 108

50-60 years: 28% = 108

Over 60 years: 9% = 34

The required sub-sample sizes were estimated for T cell, neutralizing antibody studies, antibody responses to SARS-CoV-2 variants of concern studies were based on the age categories by considering 20% (n=77) from the sample size of 384 using

20-30 years: 15% = 12

30-40 years: 20% = 15

40-50 years: 27% = 20

50-60 years: 23% = 18

60-65 years: 15% = 12

However, in the present study we managed to collect data from 437 respondents in total and out of them 80 were chosen for the T-cell study using convenience sampling.

Haemagglutination tests for detection of antibodies to the RBD in WT and SARS-CoV-2 variants

Briefly, sera were doubling-diluted in 50ul PBS in V bottomed 96 well plates, 50ul of ~1% v/v O-ve red cells were added, followed by 50ul of the relevant IH4-RBD reagent diluted to 2ug/ml (100ng/well). Plates were incubated for 1 hour at RT, tilted for ~20 seconds to allow a red cell “teardrop” to form, photographed and read by eye. The RBD-specific antibody titre for the serum sample was defined by the last well in which the complete absence of “teardrop” formation was observed. The HAT titration was performed using 7 doubling dilutions of serum from 1:20 to 1:1280, to determine presence of RBD-specific antibodies. A titre of 1:20 was considered as a positive response, as previously determined by us^2^. The IH4-RBD reagents for each VOC were standardized by titration with the monoclonal antibody EY-6A^3,4^ that binds to a conserved epitope common to all variants. A 20ug/ml solution of EY-6A titrated equally with a standard (2ug/ml) solution of each of the new IH4-RBD reagents. All were therefore added as 50ul from a 2ug/ml stock solution (100ng/well) as described^4^.

Ex vivo and cultured ELISpot assays

Ex vivo IFNγ ELISpot assays were carried out using freshly isolated peripheral blood mononuclear cells (PBMC). Individuals for T cell assays were randomly recruited from the study participants, and we included those who consented to provide an additional blood volume for T cell assays (7ml), in addition to the antibody assays (5ml). For cultures ELISpot assays, PBMC from each donor were incubated with the peptides covering the whole spike protein (253 overlapping peptides). Briefly, 5.0×10^6^ PBMCs were incubated for 10 days with 200µl of 40µM peptide pool in a 24 well plate. IL-2 was added on day 3 and 7 at a concentration of 100units/ml. All cell lines were routinely maintained in RPMI 1640 supplemented with 2mM L-glutamine, 100IU/ml penicillin and 100µg/ml plus 10% human serum at 37ºC, in 5% CO_2_^5^_._

For ex vivo and cultured ELISpot assays, two pools of overlapping peptides named S1 (peptide 1 to 130) and S2 (peptide 131 to 253) covering the whole spike protein (253 overlapping peptides) were added at a final concentration of 10 µM and incubated overnight as previously described ^6,7^. For ex vivo ELISpot assays, 100,000 cells/well were added, while for cultured ELISpot assays, 40,000 cells/well were added. All peptide sequences were derived from the wild-type consensus and were tested in duplicate. PHA was included as a positive control of cytokine stimulation and media alone was applied to the PBMCs as a negative control. An example of a ex vivo ELISpot assay for S1, S2, is shown in supplementary methods figure 1 and for the cultured ELISpot assays, in supplementary methods figure 2.

Briefly, ELISpot plates (Millipore Corp., Bedford, USA) were coated with anti-human IFNγ antibody overnight (Mabtech, Sweden). The plates were incubated overnight at 37°C and 5% CO_2_. The cells were removed, and the plates developed with a second biotinylated Ab to human IFNγ and washed a further six times. The plates were developed with streptavidin-alkaline phosphatase (Mabtech AB) and colorimetric substrate. The spots were enumerated using an automated ELISpot reader (AID Germany). Background (PBMCs plus media alone) was subtracted and data expressed as number of spot-forming units (SFU) per 10^6^ PBMCs. A positive response was defined as mean±2 SD of the background responses.

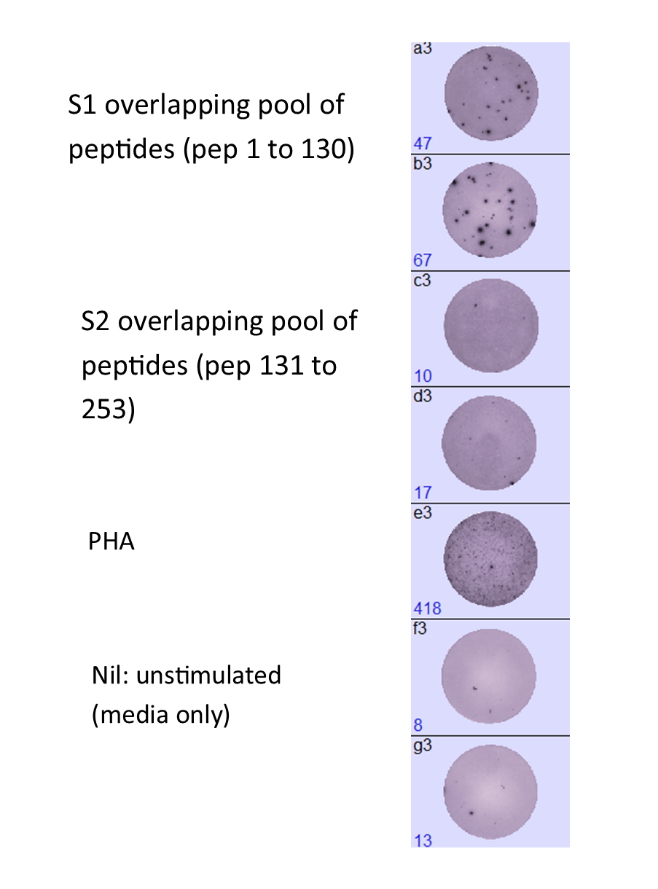

**Supplementary methods figure 1**: An example of an ex vivo ELISpot response at 6 weeks (2 weeks following the second dose of the vaccine).

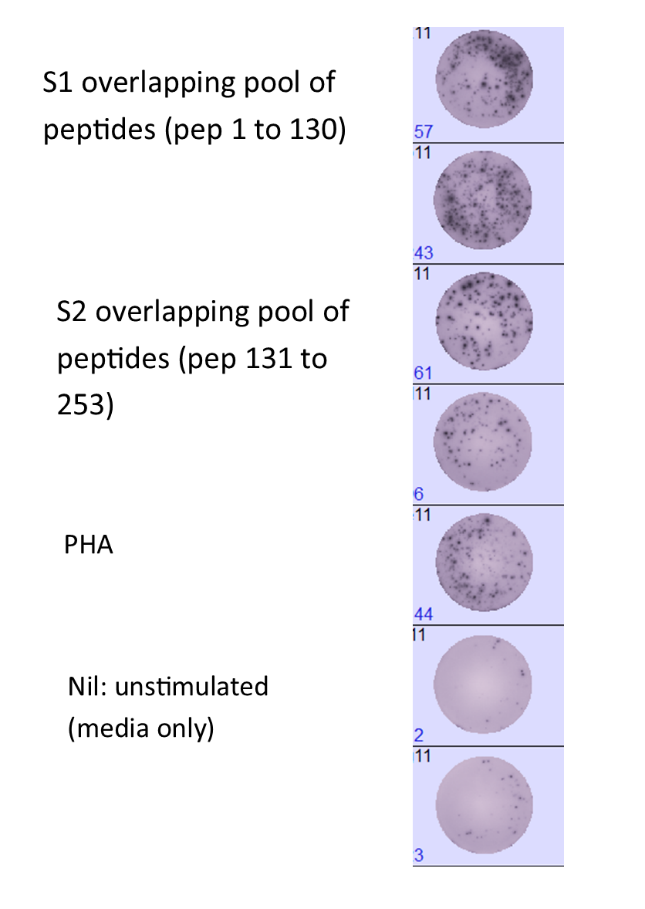

**Supplementary methods figure 2**: An example of a cultured ELISpot response at 6 weeks (2 weeks following the second dose of the vaccine).

Intracellular cytokine staining

Intracellular cytokine staining was carried out in freshly isolated PBMCs. PBMCs were incubated with CD107a FITC (Biolegend, USA) for 30 minutes in RPMI 1640 and 10% heat inactivated human serum (Sigma Andrich). Cells were stimulated with overlapping peptides pool of SARS-CoV-2 spike protein for 2 hours at 1mM concertation before adding monensin (Biolegend, USA). The PBMC were incubated for a further 14 hours before staining with with anti‐CD3 APC Cy7 (clone OKT3), anti‐CD8 BV650 (clone SK1) and anti-CD4 PB (clone OKT4). Then the cells were fixed with fixation buffer and permeabilized with perm wash buffer (Biolegend, USA) and stained for IFN-γ APC (clone 4S. B3).  Live/Dead fixable aqua dead cell stain (Thermo Fisher Scientific, USA) was used according to the manufacturer's protocol to exclude dead cells. Cells were acquired on a BD FACSAria III Cell Sorter using DIVA v8 software (BD Biosciences, USA). For each donor, unstimulated cells were included as a negative control. Flow cytometry data were analyzed using FlowJo v.10.7.1 software (FlowJo). Fluoresces Minus One (FMO) controls were used to draw the gates for both CD107a and IFN-γ (supplementary methods figure 3).

The proportion of cells expressing S pool of peptide specific CD107a or producing IFNγ, was determined by subtracting the expression levels/production levels in the unstimulated wells, from the peptide stimulated wells.

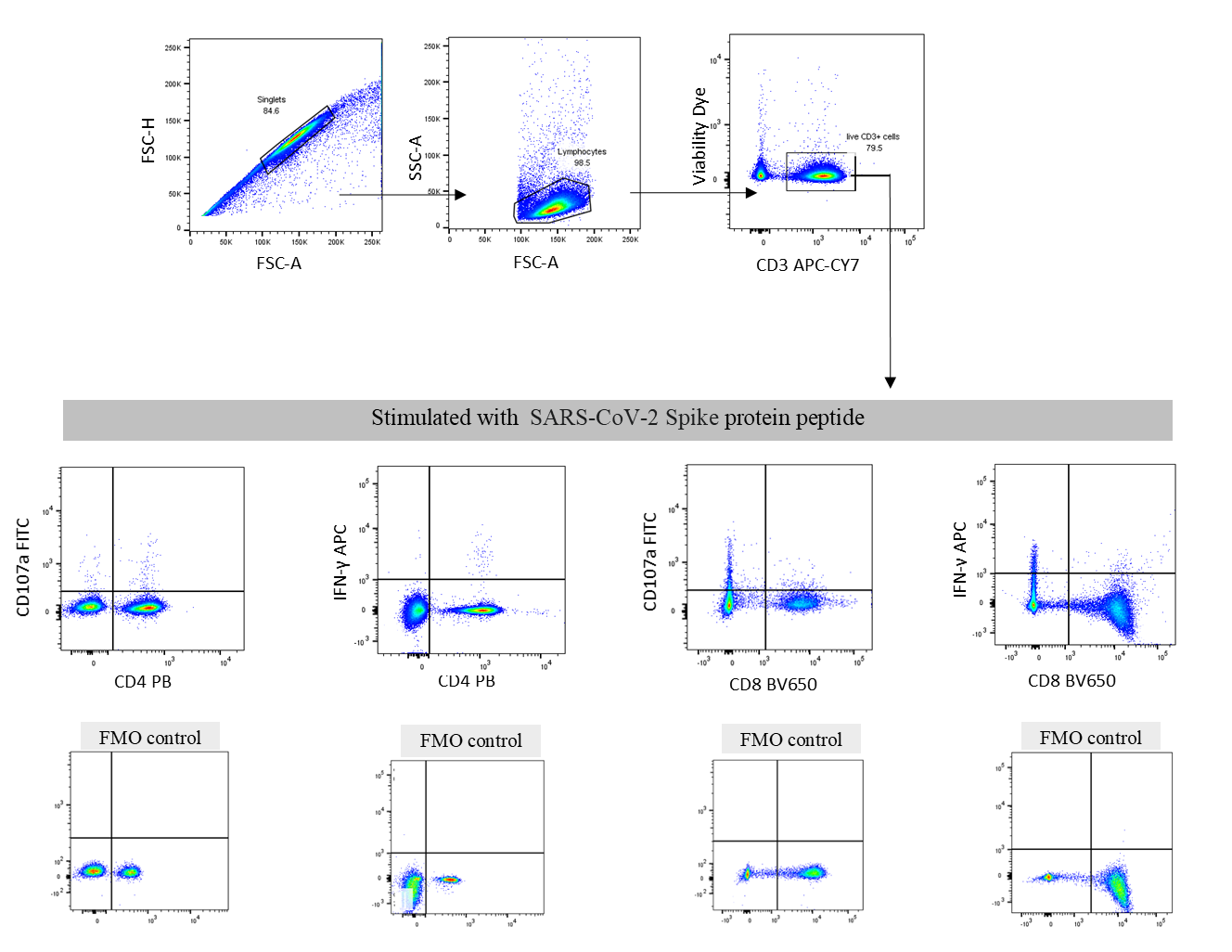

**Supplementary methods figure 3:** Gating strategy used to identify CD107a expressing CD4+ and CD8+ T cells and IFNγ producing CD4+ and CD8+ T cells. The cells were initially gates on FSC-H and FSC-A to gate the singlets. The lymphocytes from these cells were then identified by gating them on the FSC and SSC. From these cells, the live cells were then gated and then CD3+ T cells were gated. From these CD3+ T cells, CD107a expressing CD4+ and CD8+ T cells were identified, and IFNγ producing CD4+ T cells and CD8+ T cells were identified.

B cell ELISpot assays

Briefly, freshly isolated PBMCs were stimulated in a 24 well plate using IL-2 and R848 (a TLR 7/8 agonist) in RPMI supplemented with 10% fetal bovine serum, 1% penicillin streptomycin and 1% glutamine at 4 million cells/well and incubated at 37 °C with 5% CO_2_ for 3 days. They were then washed and rested overnight and 100,000 cells/well were added. 50,000 cells/well were added to the positive control wells. A Human IgG ELISpot kit (Mabtech 3850-2A) was used according to the manufacturer’s instructions to quantify IgG-secreting cells specific to SARS-COV2 S1, S2 and N recombinant proteins, which were coated at 2µg/ml in phosphate buffered saline (PBS). All experiments were carried out in duplicate and anti-human IgG monoclonal capture antibodies, was used as a positive control, and media alone as a negative control. The spots were enumerated using an automated ELISpot reader (AID Germany). A positive response was defined as mean±2 SD of the background responses. An example of a B cell ELISpot assay for S1, S2 and N recombinant proteins is shown in supplementary methods figure 4.

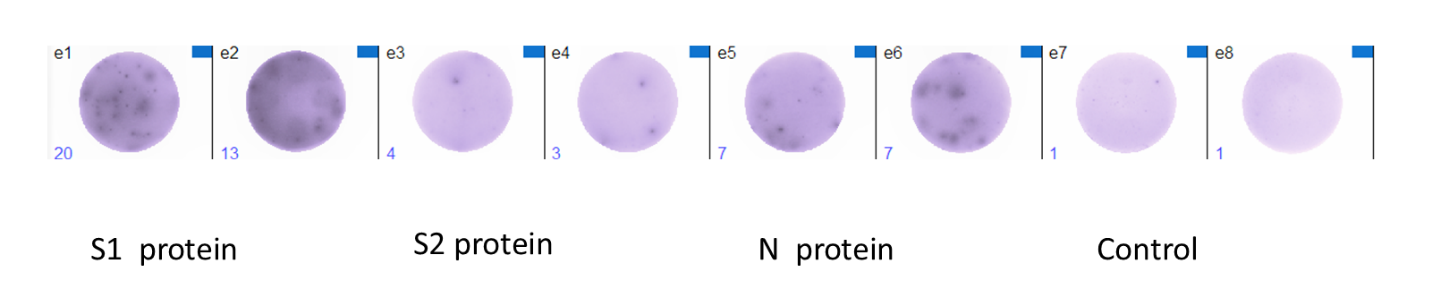

**Supplementary methods figure 4:** An example of a B cell ELISpot assay at 6 weeks (2 weeks since administering the second dose) for S1, S2 and N recombinant protein.
